# Sociodemographic and occupational factors associated with general sick leave among public school teachers in Bogotá: A retrospective cohort study, 2010–2025

**DOI:** 10.64898/2026.08.06.26359845

**Authors:** Hernando Bayona-Rodríguez, Daniela Sánchez-Santiesteban, Giancarlo Buitrago

**Affiliations:** Facultad de Ciencias Económicas, Universidad Nacional de Colombia, Bogotá, Colombia; Departamento de Epidemiología Clínica y Bioestadística, Facultad de Medicina, Universidad Nacional de Colombia, Bogotá, Colombia; Fundación Cardioinfantil - Instituto de Cardiología, Bogotá, Colombia

**Keywords:** Teachers, occupational health, Low and Middle Income Countries

## Abstract

**Background:** General illness–related sick leave among teachers represents a relevant public health and workforce management issue. However, long-term population-based evidence describing its distribution and associated factors in Latin American urban educational systems remains limited. This study aimed to characterize the occurrence, distribution, duration, and sociodemographic, occupational, temporal, and territorial factors associated with general illness–related sick leave among public school teachers in Bogotá between 2010 and 2025.

**Methods:** A retrospective cohort study was conducted using integrated administrative databases from the Bogotá District Department of Education. The primary outcome was the occurrence of at least one general illness–related sick leave episode in a teacher-month observation. Descriptive analyses were performed to characterize sociodemographic and occupational patterns. A multivariable logistic regression model was used to estimate associations. Month and year were included as temporal fixed effects to account for seasonal patterns, academic-calendar effects, pandemic-related disruption, and secular changes.

**Results:** The cohort included 59,697 unique teachers, contributing 537,025 teacher-year observations from teachers with an active employment record between January 1, 2010, and July 31, 2025. Overall, 41.59% of teacher-year observations included at least one general illness–related sick leave episode, and 83.26% of teachers had at least one episode at any time during follow-up. Respiratory diseases accounted for the largest share of episodes, followed by musculoskeletal and infectious diseases. Mean duration varied substantially by diagnostic category, ranging from short respiratory and infectious episodes to longer absences related to neoplasms, circulatory diseases, injuries, and mental health conditions. In the multivariable teacher-month model, sick leave occurrence was associated with age, sex, occupational role, teaching area, contract type, locality, calendar month, and calendar year. Lower odds were observed among male teachers, principals, and teachers with provisional contracts, while temporal and territorial variation was observed across months, years, and localities.

**Conclusions:** General illness–related sick leave among public school teachers in Bogotá showed consistent sociodemographic, occupational, temporal, and territorial patterns. Respiratory and musculoskeletal conditions accounted for the largest share of episodes, while chronic, neoplastic, injury-related, circulatory, and mental health conditions were associated with longer durations. These findings provide population-level evidence to inform occupational health surveillance, seasonal preparedness, and workforce planning strategies within urban educational systems.

## Introduction

Medically certified sick leave among teachers represents a complex and multifactorial occupational health issue affecting educational systems across diverse contexts and levels of development [1]. Its consequences extend beyond individual morbidity, affecting teachers’ well-being, school organization, continuity of instruction, replacement planning, administrative costs, and the stability of educational services [2,3]. Regular teacher presence in the classroom is essential for consolidating strong educational systems, given its central role in citizenship formation, equity promotion, and human capital development [4,5]. Recurrent absences not only increase adverse health outcomes among teachers but also reduce effective instructional time, generate additional administrative burden, and weaken the quality and continuity of the educational process [6–8]. In this context, advancing the understanding of determinants of teacher sick leave is essential to inform evidence-based prevention and mitigation strategies.

Teaching is an occupation with distinctive health-related demands. Teachers are exposed to sustained vocal use, prolonged standing, repetitive postures, high cognitive and emotional demands, psychosocial stressors, and close interpersonal contact with students and colleagues. These conditions may contribute to respiratory, musculoskeletal, vocal, and mental health problems, which have been frequently described as relevant causes of morbidity and work absence in this occupational group [9,10]. In addition, teacher sick leave may vary according to sociodemographic and occupational characteristics such as sex, age, occupational role, employment conditions, school context, and temporal factors related to the academic calendar [1].

Most available evidence on teacher absenteeism in Latin America has focused on general absence from school, working conditions, or educational system performance [11,12]. Although this literature is valuable, it does not always distinguish medically certified sick leave from other forms of absence, nor does it consistently use longitudinal individual-level administrative data. In Colombia, previous evidence on sick leave among teachers has reported sex differences and identified musculoskeletal and respiratory conditions among the most frequent causes of medically certified absence [13–15]. However, limited population-based evidence is available on the annual occurrence of general illness–related sick leave, its diagnostic profile, duration, and associated sociodemographic and occupational factors in large urban teaching workforces in the region.

Colombia provides a relevant setting for studying this issue. The education system is characterized by high workload demands, limited resources, and frequent sociopolitical pressures, conditions that may intensify occupational risks and health inequalities among teachers [14,16]. National absence statistics also suggest a substantial burden of work absence; for instance, in 2022, an average of 11.8 days of work absence per worker per year was reported according to data from the National Business Association, including common illness, leave, and permits. Within the public teaching workforce, previous evidence from Bogotá identified female sex as a factor associated with a higher likelihood of teacher sick leave and reported musculoskeletal and respiratory conditions among the most frequent causes [17]. Public school teachers in Colombia operate under a special social security regime, the National Fund for Social Benefits of the Teaching Profession (FOMAG, for its acronym in Spanish), which enables standardized administrative recording of health-related leave events. In Bogotá, the country’s largest urban educational system, these records offer an opportunity to examine general illness–related sick leave using population-level administrative data rather than surveys, selected institutions, or limited samples.

Recently, our group reported national evidence on the health of Colombian teachers using administrative databases from the contributory health insurance scheme. That study focused on private-sector teachers and evaluated mortality and selected comorbidities during a single calendar year, demonstrating that teachers exhibited a different health profile compared with other workers in the same occupational risk category [18]. However, it did not evaluate medically certified sick leave or include public school teachers covered by Colombia’s special teachers’ social security regime (FOMAG). Consequently, important evidence gaps remain regarding the occurrence, duration, and determinants of illness-related absenteeism among public school teachers, who represent a distinct workforce with different employment conditions and healthcare organization.

The present study aimed to characterize the occurrence, diagnostic distribution, duration, and sociodemographic, occupational, temporal, and territorial factors associated with general illness–related sick leave among public school teachers in Bogotá between 2010 and 2025. In the Colombian context, this workforce operates under a special social security regime, the FOMAG, which enables standardized administrative recording of health-related leave events. To our knowledge, this is the first population-based study to examine general illness–related sick leave among public school teachers in Bogotá using administrative records from the public education system. Bogotá is a highly heterogeneous urban setting, with marked differences across localities in socioeconomic conditions, environmental exposures, school infrastructure, and access to services. Although these contextual factors were not directly measured in this study, they provide a relevant rationale for examining territorial variation in sick leave patterns. This approach provides quantitative evidence on the magnitude, distribution, and factors associated with general illness–related sick leave and may support occupational health surveillance, teacher workforce planning, and sustainable absence coverage strategies in the public education system.

## Methods

### Ethical considerations

This study was granted institutional review board (IRB) ethical approval by the Institutional Ethics Committee of Facultad de Ciencias Económicas at Universidad Nacional de Colombia (Approval Number: B.FCE.1.002-518-25). Written consent was waived by the IRB as data sources were administrative databases fully anonymised. Following ethics approval, initial access to the databases was on 15/10/2025 with the purpose of identifying the population and creating a refined dataset for subsequent analysis. This refined dataset forms the basis of the analyses presented in this paper.

### Study design and population

A retrospective cohort study was conducted using administrative databases from the Bogotá District Department of Education (Secretaría de Educación del Distrito, SED), Colombia. The study population included teachers, academic coordinators, directors, and school principals formally employed within the district teaching workforce under either permanent or provisional contracts and assigned to an official public educational institution during the study period. The descriptive observation period spanned from January 1, 2010, to July 31, 2025. For the main multivariable analysis, data were structured as teacher-month observations, with each individual contributing one observation for each calendar month in which they had an active employment record. Records with missing key sociodemographic or occupational information (sex, age, employment type, or position) were excluded.

### Data and materials

The data sources for this study comprised three administrative databases provided by the Bogotá District Department of Education (SED) (Additional File 1). The first database corresponded to the teacher workforce registry, which includes the employee identification code, workplace characteristics (locality, school code, administrative unit, work schedule, educational level, teaching area, employment type, position, and year), as well as sociodemographic variables such as sex and date of birth. The second database was the sick leave registry from 2010–2017, which records sick leave episodes during the 2010–2017 period. This database includes teacher identification variables (employee code, enabling linkage with the workforce registry) and variables related to sick leave episodes, such as start and end dates, number of days, and type of leave. Finally, a third sick leave database was used, covering the period from 2018 onward. This registry includes the same variables described for the previous database and additionally incorporates the ICD-10 diagnostic code associated with each sick leave episode, according to the International Classification of Diseases.

### Variables

The primary outcome was a binary indicator of whether a teacher-month observation included at least one episode of general illness–related sick leave during that calendar month. General illness–related sick leave was defined according to the administrative leave type recorded in the sick leave registry. Maternity and paternity leave were recorded as separate administrative categories and were therefore not included in this outcome. General illness–related episodes were linked to the corresponding teacher-month observation using the employee identification code and the calendar month and year of the episode start date. If more than one general illness–related sick leave episode occurred within the same teacher-month, the primary outcome remained coded as one. For descriptive purposes, teacher-month observations were also aggregated at the teacher-year level to estimate the annual proportion of teachers with at least one general illness–related sick leave episode. Sick leave episodes were analyzed separately to describe diagnostic distribution and duration.

Baseline variables included sociodemographic and occupational characteristics. Sociodemographic variables comprised sex (female or male) and age, calculated as of January 1 of each calendar year with an active employment record. Occupational variables included the recorded position (teacher, coordinator, director, or principal), the educational level in which the individual worked (preschool, primary education, lower secondary education, upper secondary education, complementary cycle, or global assignment), teaching area (mathematics; biology and chemistry; technology; higher education; social sciences; philosophy; language—Spanish or foreign language; arts and cultural education; physical education and health; religious and ethics education; or special and differentiated education), work schedule (full-time or extended, weekend, intermediate, morning, night, afternoon, or single shift), type of contract (permanent or provisional), and the locality where the educational institution was located.

### Analysis

A descriptive analysis of the sociodemographic and occupational characteristics of both sick leave records and teachers was performed. Categorical variables were summarized using absolute frequencies and proportions. Continuous numerical variables were evaluated according to their distribution: variables with symmetric distribution were presented as mean and standard deviation, whereas variables with asymmetric distribution were reported as median (p50) and interquartile range (p25–p75). Population analyses were conducted by stratifying according to year of employment and according to the presence or absence of sick leave during the observation period. Descriptive analyses included all available data from 2010 through July 31, 2025. Annual descriptive summaries were based on teacher-year observations and were used to estimate the proportion of teachers with at least one general illness–related sick leave episode during each calendar year. Diagnostic distribution and duration analyses were conducted at the sick leave episode level. Because ICD-10 diagnostic information was available only from 2018 onward, diagnostic analyses were restricted to the 2018–2025 period.

A multivariable logistic regression model was fitted to identify sociodemographic, occupational, territorial, and temporal factors associated with the occurrence of at least one general illness–related sick leave episode in a teacher-month observation. The model included sex, age, position, educational level, teaching area, work schedule, employment type, locality, calendar month, and calendar year as covariates. Calendar month and calendar year were included as temporal fixed effects to account for seasonal patterns, academic-calendar effects, pandemic-related disruption, and secular changes over the study period. Because a single individual could contribute repeated monthly observations over several years, standard errors were clustered at the individual level to account for within-person correlation over time. All multivariable regression analyses were restricted to complete calendar years from January 1, 2010, to December 31, 2024. Estimates were reported as odds ratios with 95% confidence intervals. All analyses were conducted using Stata version 18.5 MP [19]. This article followed the STROBE (Strengthening the Reporting of Observational Studies in Epidemiology) guidelines to ensure transparency and thoroughness in reporting the findings (Additional File, 2) [20].

## Results

Between January 1, 2010, and July 31, 2025, a total of 537,754 teacher-year records from the Bogotá public education workforce were identified. After excluding [XX] records with incomplete key sociodemographic or occupational information, the annual descriptive dataset included 537,025 teacher-year records corresponding to 59,697 unique individuals in the open cohort. For the main multivariable analysis, the dataset was restructured into teacher-month observations and restricted to complete calendar years from January 2010 to December 2024 (Fig 1).

**Fig 1.**
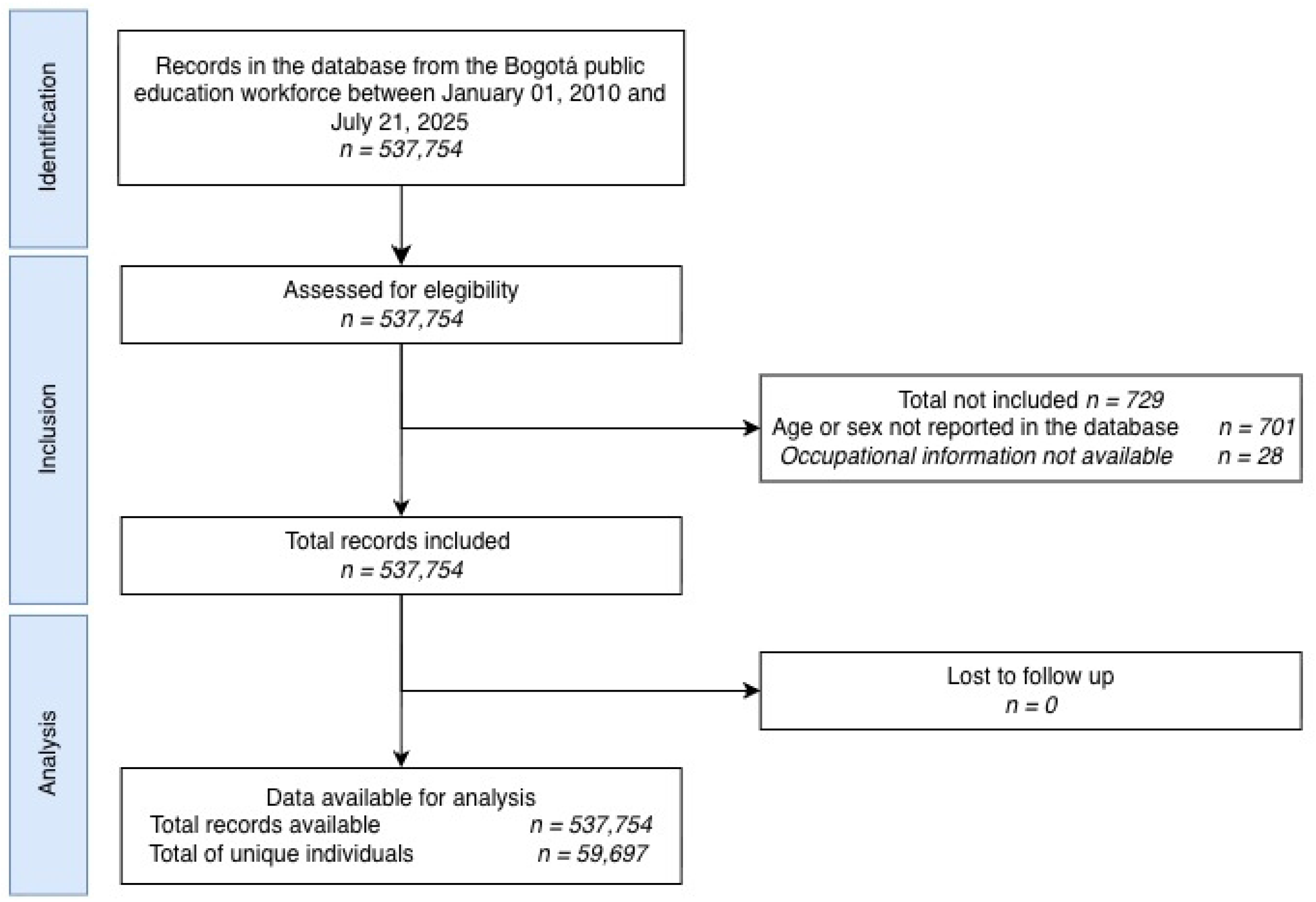
Identification of population.

Among the sociodemographic characteristics, 69.38% of the workforce were female (n = 372,598), and the mean age was 46.32 years (SD 10.61). Regarding occupational characteristics, 94.64% held teaching positions (n = 508,218), followed by coordinators (4.32%, n = 23,176). The most frequent educational level was lower and upper secondary education (47.16%, n = 253,261), followed by primary education (32.30%, n = 173,464) and preschool (10.45%, n = 56,096).

By teaching area, the largest proportion corresponded to primary education fields (26.94%, n = 144,684), followed by language (Spanish and foreign languages) (12.06%, n = 64,782) and preschool (9.77%, n = 52,469). With respect to work schedule, 43.84% worked in the morning shift (n = 235,436) and 39.45% in the afternoon shift (n = 211,849). Permanent contracts accounted for 84.47% of employment arrangements (n = 453,644). The localities with the highest concentration of teachers were Kennedy (13.59%, n = 72,960), Ciudad Bolívar (11.31%, n = 60,735), and Bosa (11.24%, n = 60,367).

During the observation period, 223,336 teacher-year observations included at least one general illness–related sick leave episode, corresponding to 41.59% of all teacher-year observations. Analyses of selected calendar years (2010, 2015, 2020, and 2025) showed broadly similar sociodemographic and occupational distributions among active public teaching workforce records over time (Table 1). In all selected years, women represented more than 68% of active records. Mean age increased from 44.53 years (SD 10.18) in 2010 to 48.51 years (SD 11.66) in 2025. In each selected year, more than 90% of records corresponded to teaching positions. Lower and upper secondary education remained the most frequent educational level, followed by primary education and preschool.

**Table 1.**
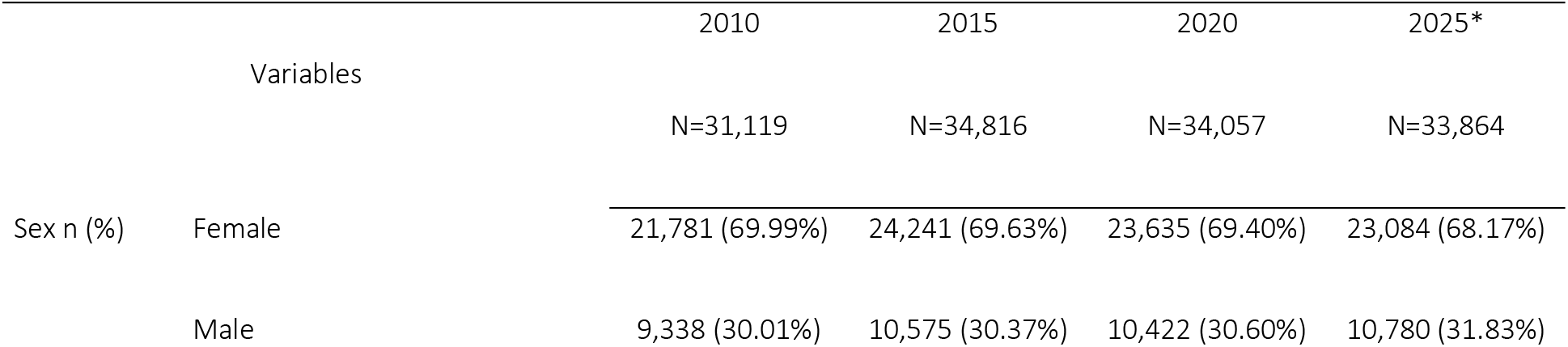
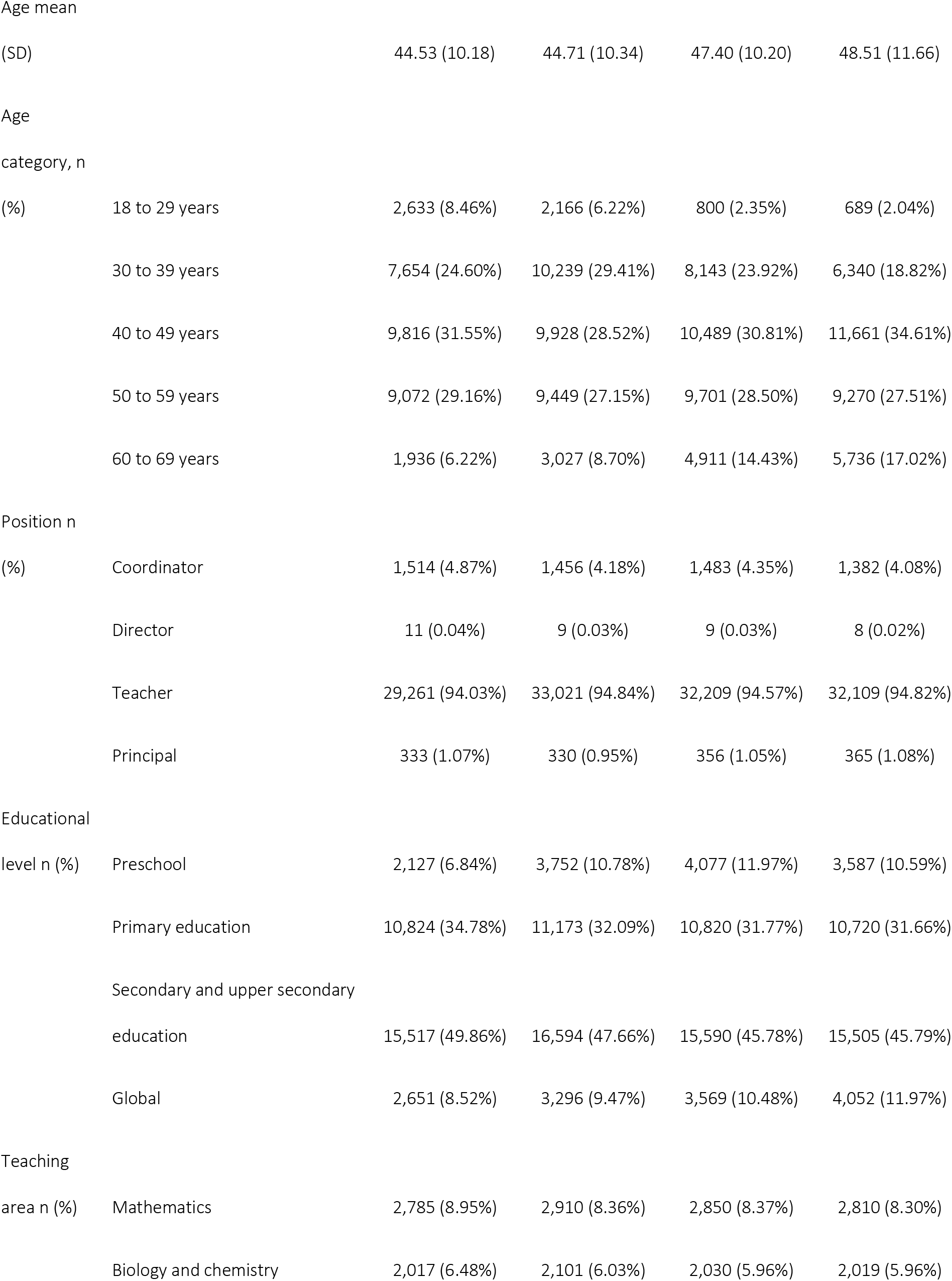
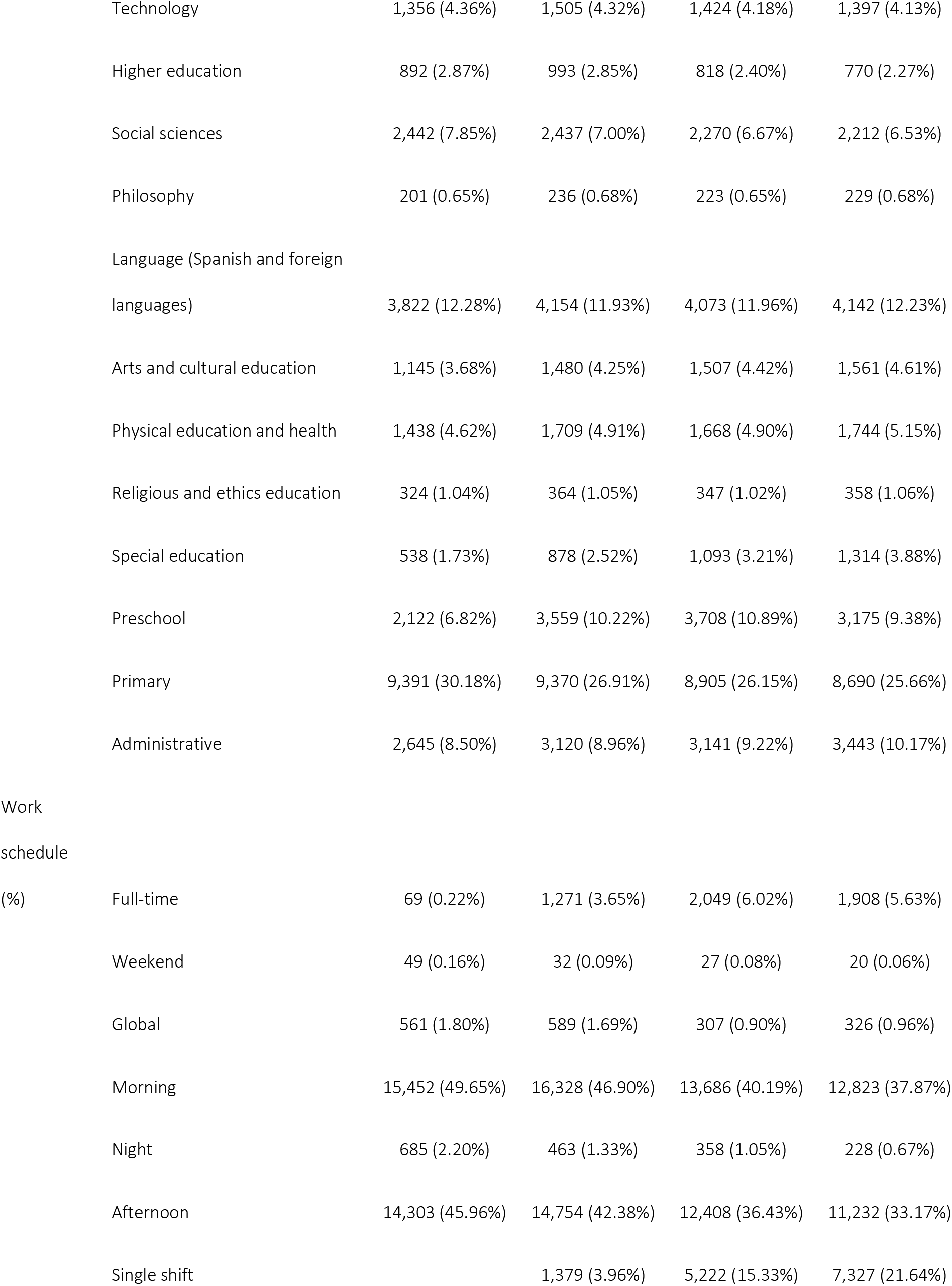
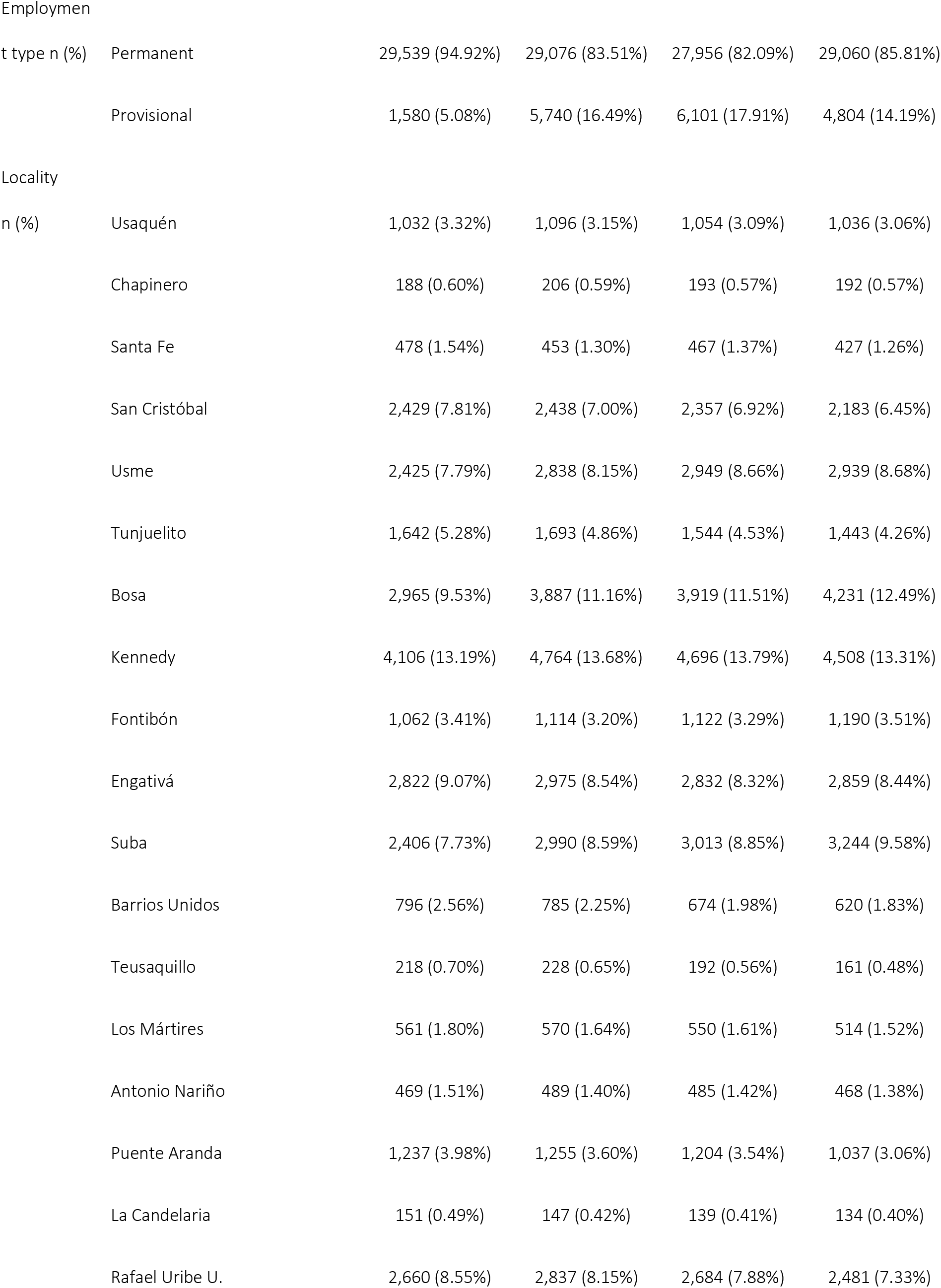
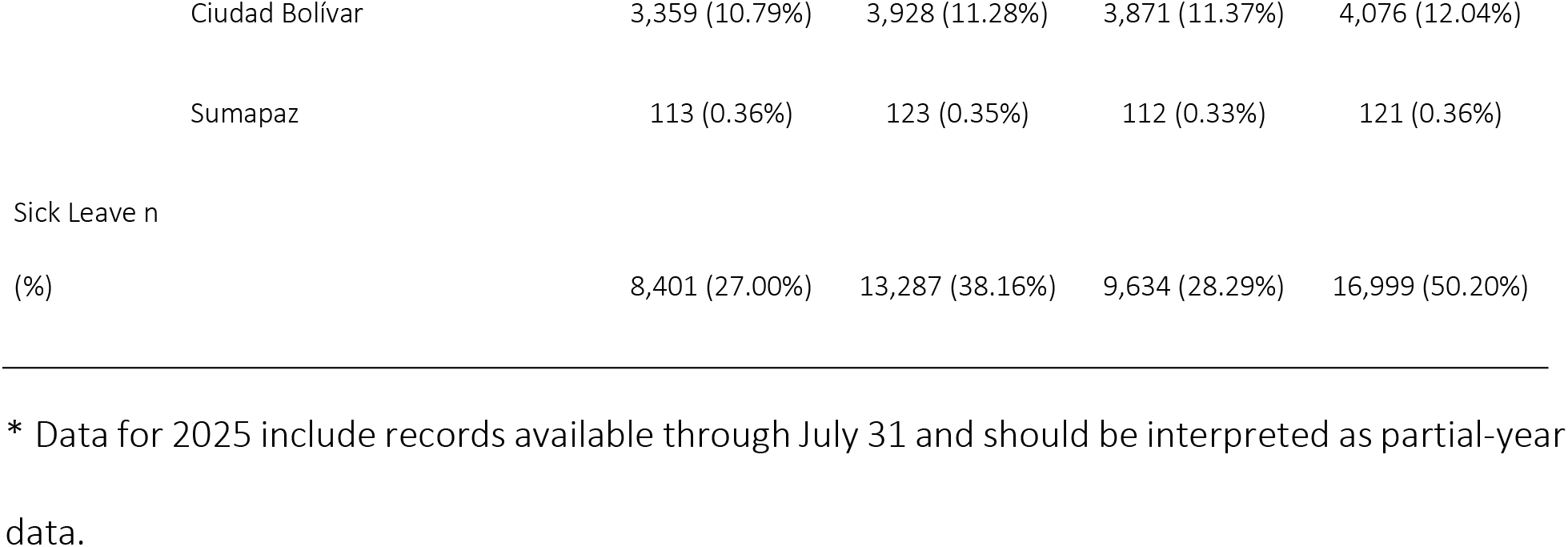
Sociodemographic and occupational characteristics of active public teaching workforce records in selected calendar years, 2010, 2015, 2020, and 2025.

Teaching areas associated with primary education, language, and preschool represented the largest shares of active records across the selected years. More than 70% of records corresponded to morning or afternoon shifts, and most employment arrangements were permanent contracts. Kennedy, Ciudad Bolívar, and Bosa consistently had the highest concentration of active public teaching workforce records. The annual proportion of records with at least one general illness–related sick leave episode increased from 27.00% in 2010 to 38.16% in 2015 and 50.20% in 2025, based on partial-year data available through July 31. In 2020, the corresponding proportion was 28.29%. This decrease occurred during the first year of the COVID-19 pandemic, when remote work and school closures may have changed exposure patterns and sick leave certification practices.

Throughout the study period, general illness–related sick leave accounted for the largest proportion of registered sick leave episodes, followed by occupational disease and, to a lesser extent, maternity or paternity leave. Overall, 88.83% of all registered episodes corresponded to general illness (n = 514,277), 9.40% to occupational disease (n=54,428), 1.27% to maternity or paternity leave (n = 7,377), and 0.50% to prolonged sick leave exceeding 90 days (n=2,875) (Fig 2).

**Fig 2.**
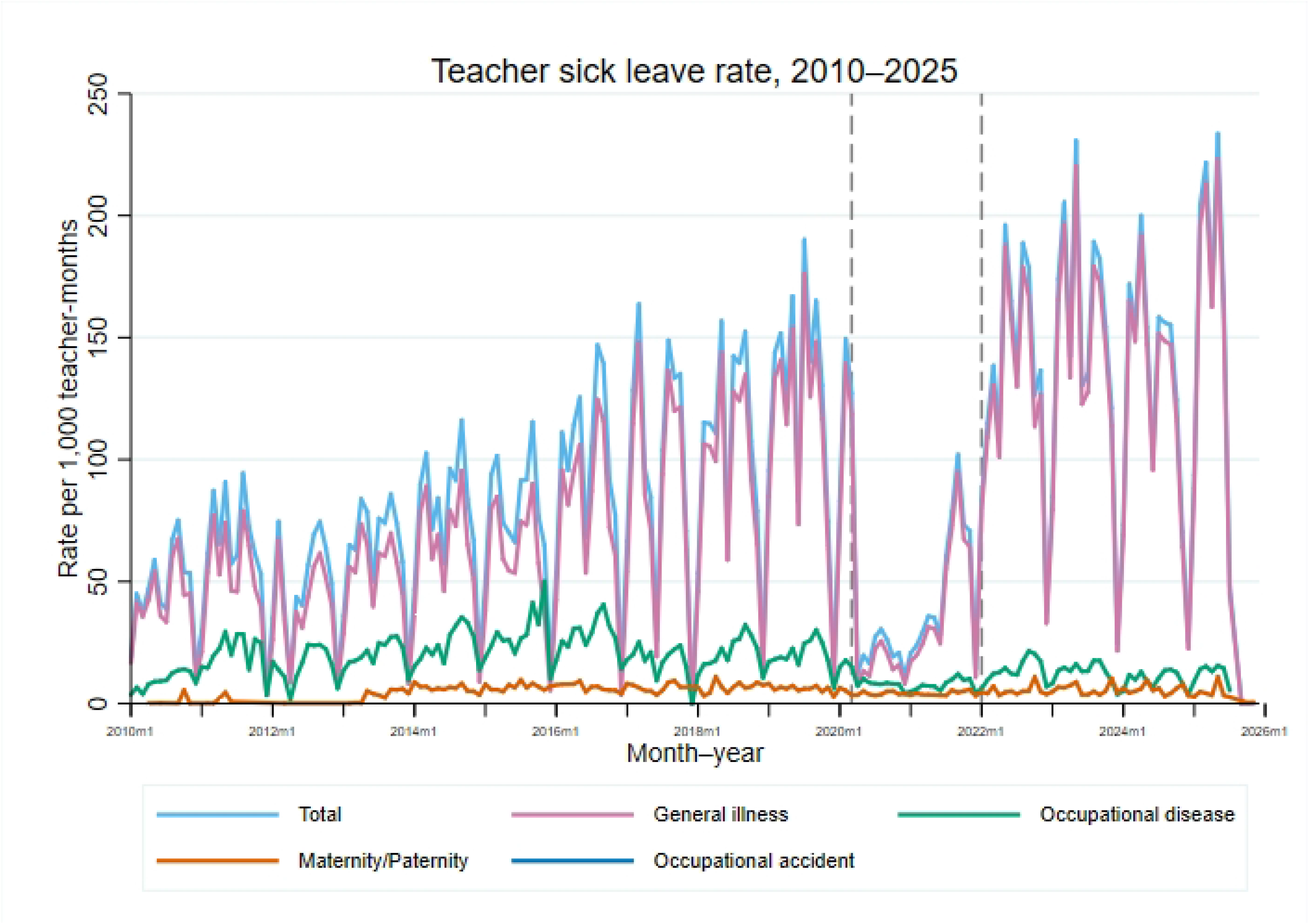
Annual rate of registered teacher sick leave episodes by type of leave, 2010–2025.

Among general illness–related sick leave episodes with available ICD-10 diagnostic information during the 2018–2025 period, respiratory diseases represented the largest diagnostic category (27.68%; n = 89,864), followed by musculoskeletal and connective tissue diseases (10.45%; n = 33,919) and infectious and parasitic diseases (9.93%; n = 32,230) (Table 2). The remaining diagnostic categories each accounted for less than 7% of episodes.

**Table 2.** Distribution of general illness–related teacher sick leave episodes according to ICD-10 diagnostic categories, 2018–2025.

| ICD-10 diagnostic category | Frequency | Percentage |
| --- | --- | --- |
| Respiratory diseases | 89,864 | 27.68% |
| Musculoskeletal and connective tissue diseases | 33,919 | 10.45% |
| Infectious and parasitic diseases | 32,230 | 9.93% |
| Digestive system diseases | 21,603 | 6.65% |
| Injury, poisoning, and external causes | 20,651 | 6.36% |
| Factors influencing health status and contact with health services | 18,872 | 5.81% |
| Diseases of the central nervous system | 14,811 | 4.56% |
| Diseases of the eye and ear | 13,642 | 4.20% |
| Genitourinary system diseases | 11,521 | 3.55% |
| Mental and behavioral disorders | 11,217 | 3.46% |

The mean duration of general illness–related sick leave varied substantially across diagnostic categories. Neoplasms and haematologic diseases had the longest mean duration (18.7 days; 95% CI 18.39–18.95), followed by congenital malformations and chromosomal abnormalities (14.2 days; 95% CI 12.54–15.94) and perinatal conditions (13.3 days; 95% CI 12.01–14.68). Categories with intermediate mean durations included circulatory system diseases, injury and other external causes, pregnancy, childbirth and the puerperium, and mental and behavioural disorders. By contrast, respiratory diseases and infectious and parasitic diseases had the shortest mean durations, at approximately 2.4 days each.

In the multivariable logistic regression model based on teacher-month observations from complete calendar years between 2010 and 2024, age was positively associated with the monthly occurrence of at least one general illness–related sick leave episode (OR 1.003; 95% CI 1.002–1.005) (Table 3). Compared with female teachers, male teachers had lower odds of monthly sick leave occurrence (OR 0.538; 95% CI 0.523–0.554). Using coordinators as the reference category, teachers had higher odds of sick leave (OR 1.351; 95% CI 1.271–1.435), whereas principals had lower odds (OR 0.452; 95% CI 0.384–0.533). Regarding teaching area, with mathematics as the reference category, higher odds of monthly sick leave occurrence were observed among teachers in arts and cultural education (OR 1.121; 95% CI 1.043–1.205) and physical education and health (OR 1.124; 95% CI 1.047–1.207). Lower odds were observed in special and differentiated education (OR 0.863; 95% CI 0.791–0.942) and preschool education (OR 0.682; 95% CI 0.612–0.759). Teachers with provisional contracts had lower odds of monthly sick leave occurrence compared with those holding permanent contracts (OR 0.619; 95% CI 0.601–0.638).

**Table 3.** Multivariable logistic regression analysis of sociodemographic, occupational, territorial, and temporal factors associated with monthly general illness–related teacher sick leave, 2010–2024.

| Variable | OR |  | IC 95% | p-value |
| --- | --- | --- | --- | --- |
| Age | 1.003 | 1.002 | 1.005 | <0.001 |
| Sex |  |  |  |  |
| Female | Ref |  |  |  |
| Male | 0.538 | 0.523 | 0.554 | <0.001 |
| Position |  |  |  |  |
| Coordinator | Ref |  |  |  |
| Director | 0.603 | 0.248 | 1.467 | 0.265 |
| Teacher | 1.351 | 1.271 | 1.435 | <0.001 |
| Principal | 0.452 | 0.384 | 0.533 | <0.001 |
| Educational level |  |  |  |  |
| Preschool | Ref |  |  |  |
| Primary education | 0.904 | 0.879 | 0.930 | <0.001 |
| Secondary and upper secondary education | 0.766 | 0.745 | 0.789 | <0.001 |
| Global | 0.708 | 0.683 | 0.734 | <0.001 |
| Teaching area |  |  |  |  |
| Mathematics | Ref |  |  |  |
| Biology and chemistry | 0.996 | 0.933 | 1.062 | 0.898 |
| Technology | 1.005 | 0.934 | 1.082 | 0.888 |
| Higher education | 0.988 | 0.909 | 1.073 | 0.769 |
| Social sciences | 1.012 | 0.950 | 1.077 | 0.718 |
| Philosophy | 0.960 | 0.839 | 1.098 | 0.548 |
| Language (Spanish and foreign languages) | 1.003 | 0.950 | 1.060 | 0.903 |
| Arts and cultural education | 1.121 | 1.043 | 1.205 | 0.002 |
| Physical education | 1.124 | 1.047 | 1.207 | 0.001 |
| Religious and ethics education | 0.989 | 0.869 | 1.126 | 0.871 |
| Special and differentiated education | 0.863 | 0.791 | 0.942 | 0.001 |
| Work Schedule |  |  |  |  |
| Full time | Ref |  |  |  |
| Weekend | 1.235 | 0.892 | 1.710 | 0.204 |
| Global | 1.284 | 0.115 | 14.289 | 0.839 |
| Morning | 1.054 | 0.625 | 1.779 | 0.843 |
| Night | 1.245 | 0.882 | 1.757 | 0.214 |
| Afternoon | 0.528 | 0.189 | 1.474 | 0.223 |
| Single Shift | 1.289 | 0.931 | 1.786 | 0.126 |
| Employment type |  |  |  |  |
| Permanent | Ref |  |  |  |
| Provisional | 0.619 | 0.601 | 0.638 | <0.001 |
| Locality |  |  |  |  |
| Usaquén | Ref |  |  |  |
| Chapinero | 1.025 | 0.863 | 1.217 | 0.779 |
| Santa Fe | 0.947 | 0.855 | 1.049 | 0.295 |
| San Cristóbal | 1.025 | 0.957 | 1.098 | 0.481 |
| Usme | 0.946 | 0.884 | 1.011 | 0.102 |
| Tunjuelito | 0.974 | 0.902 | 1.051 | 0.492 |
| Bosa | 1.079 | 1.010 | 1.153 | 0.023 |
| Kennedy | 1.095 | 1.026 | 1.168 | 0.006 |
| Fontibón | 0.996 | 0.916 | 1.083 | 0.925 |
| Engativá | 1.053 | 0.983 | 1.127 | 0.140 |
| Suba | 1.134 | 1.061 | 1.211 | 0.000 |
| Barrios Unidos | 1.114 | 1.020 | 1.217 | 0.017 |
| Teusaquillo | 1.163 | 1.012 | 1.337 | 0.034 |
| Los Mártires | 0.972 | 0.881 | 1.072 | 0.571 |
| Antonio Nariño | 0.970 | 0.875 | 1.074 | 0.552 |
| Puente Aranda | 0.962 | 0.887 | 1.043 | 0.343 |
| La Candelaria | 0.911 | 0.777 | 1.068 | 0.249 |
| Rafael Uribe U. | 0.989 | 0.925 | 1.058 | 0.754 |
| Ciudad Bolívar | 1.034 | 0.969 | 1.103 | 0.316 |
| Sumapaz | 0.313 | 0.253 | 0.388 | 0.000 |
| Month |  |  |  |  |
| January | Ref |  |  |  |
| February | 2.153 | 2.116 | 2.191 | 0.000 |
| March | 2.256 | 2.216 | 2.296 | 0.000 |
| April | 1.736 | 1.704 | 1.768 | 0.000 |
| May | 2.197 | 2.159 | 2.237 | 0.000 |
| June | 1.308 | 1.283 | 1.333 | 0.000 |
| July | 2.023 | 1.987 | 2.059 | 0.000 |
| August | 2.444 | 2.401 | 2.487 | 0.000 |
| September | 2.587 | 2.541 | 2.633 | 0.000 |
| October | 1.935 | 1.901 | 1.971 | 0.000 |
| November | 1.437 | 1.411 | 1.464 | 0.000 |
| December | 0.293 | 0.285 | 0.301 | 0.000 |
| Year |  |  |  |  |
| 2010 | Ref |  |  |  |
| 2011 | 1.397 | 1.359 | 1.436 | 0.000 |
| 2012 | 1.075 | 1.043 | 1.109 | 0.000 |
| 2013 | 1.558 | 1.510 | 1.606 | 0.000 |
| 2014 | 2.103 | 2.041 | 2.168 | 0.000 |
| 2015 | 2.065 | 2.004 | 2.129 | 0.000 |
| 2016 | 2.817 | 2.733 | 2.904 | 0.000 |
| 2017 | 2.866 | 2.781 | 2.954 | 0.000 |
| 2018 | 3.138 | 3.042 | 3.236 | 0.000 |
| 2019 | 3.801 | 3.685 | 3.921 | 0.000 |
| 2020 | 1.176 | 1.134 | 1.219 | 0.000 |
| 2021 | 1.233 | 1.189 | 1.279 | 0.000 |
| 2022 | 4.170 | 4.037 | 4.307 | 0.000 |
| 2023 | 5.015 | 4.854 | 5.180 | 0.000 |
| 2024 | 4.183 | 4.048 | 4.322 | 0.000 |

Abbreviations: OR: odds ratio; CI: confidence interval. The outcome was the occurrence of at least one general illness–related sick leave episode in a teacher-month observation. Standard errors were clustered at the individual level. Calendar month and calendar year were included as temporal fixed effects.

When locality was examined using Usaquén as the reference category, higher odds of monthly sick leave occurrence were observed in Bosa, Kennedy, Suba, Barrios Unidos, and Teusaquillo. The lowest odds were observed in Sumapaz (OR 0.313; 95% CI 0.253–0.388). Temporal variation was observed across both calendar months and years. Compared with January, the odds of monthly sick leave occurrence were higher from February through November, with the highest odds observed in September (OR 2.587; 95% CI 2.541–2.633), whereas December showed lower odds (OR 0.293; 95% CI 0.285–0.301). Compared with 2010, higher odds were observed in most subsequent years, particularly in 2022 (OR 4.170; 95% CI 4.037–4.307), 2023 (OR 5.015; 95% CI 4.854–5.180), and 2024 (OR 4.183; 95% CI 4.048–4.322).

## Discussion

This study provides population-based evidence on general illness–related sick leave among public school teachers in Bogotá over a long observation period. Using integrated administrative databases from the Bogotá District Department of Education, we analyzed more than 500,000 teacher-year observations and characterized the occurrence, distribution, diagnostic profile, and temporal patterns of medically certified sick leave in the public teaching workforce. Approximately four out of ten teacher-year observations included at least one general illness–related sick leave episode, indicating a substantial annual burden of medically certified absence in this occupational group.

The findings show that general illness–related sick leave was not randomly distributed across the teaching workforce. Instead, it followed consistent sociodemographic, occupational, temporal, and territorial patterns. Older age was positively associated with the monthly occurrence of at least one general illness–related sick leave episode. This result is compatible with occupational health literature suggesting that aging workforces may experience a greater burden of health-related work absence due to accumulated morbidity, longer exposure to occupational demands, and changes in work ability over time [21,22].

Sex differences were also observed. Male teachers had lower odds of general illness–related sick leave compared with female teachers. This pattern is consistent with previous studies reporting sex-based differences in medically certified absence and occupational morbidity [23–26]. Importantly, maternity and paternity leave were not included in the general illness outcome analyzed in this study. Therefore, the observed association between sex and general illness–related sick leave is unlikely to be explained by the inclusion of administratively classified parental leave. Nevertheless, this association should not be interpreted as evidence of biological vulnerability alone. It may reflect a combination of differences in health needs, healthcare-seeking behavior, occupational exposures, and gendered social roles that influence medically certified absence.

Respiratory diseases represented the largest proportion of general illness–related sick leave episodes, followed by musculoskeletal and infectious conditions. These results are consistent with the occupational characteristics of teaching, which include prolonged indoor exposure, close interpersonal contact, sustained vocal use, and exposure to high-density classroom environments. Although respiratory conditions were the most frequent, their mean duration was among the shortest. In contrast, chronic and complex diagnoses, including neoplasms, circulatory diseases, injuries, and mental and behavioral disorders, were associated with longer absence durations. This contrast suggests that frequency and duration capture different dimensions of the sick leave burden [27–29].

The predominance of respiratory diseases has direct implications for occupational prevention in schools. Because respiratory-related sick leave may be influenced by classroom density, indoor air quality, ventilation, seasonal circulation of infectious agents, and close interaction between teachers and students, prevention should not rely exclusively on individual-level measures. Educational and occupational health authorities could consider strategies such as improving classroom ventilation, reducing overcrowding where feasible, strengthening infection-prevention practices, promoting vaccination among school staff, and facilitating timely access to medical care. In addition, adequate recovery periods during peaks of respiratory illness may help reduce presenteeism and recurrent short-term absences.

Musculoskeletal conditions were also an important component of general illness–related sick leave. Teaching involves prolonged standing, repetitive movements, sustained postures, writing on boards, carrying materials, and classroom management activities. These exposures may contribute to musculoskeletal symptoms and recurrent absence. Occupational health programs for teachers should therefore include ergonomic assessment of classrooms and work tasks, active breaks during the workday, early identification of musculoskeletal symptoms, access to rehabilitation, and adaptations for teachers with recurrent or chronic conditions.

Mental and behavioral disorders accounted for a smaller proportion of episodes but showed longer mean duration than several acute diagnostic groups. This pattern is relevant because longer absences may generate greater disruption in school organization, replacement needs, and continuity of teaching activities. In the teaching profession, psychosocial demands, emotional labor, administrative burden, classroom management, and work overload may contribute to mental health problems. These findings support the inclusion of mental health surveillance, early detection of distress, burnout prevention, workload management, and access to psychological support within teacher occupational health programs.

Temporal variation was observed across both calendar months and years. Because the primary outcome in the main model was defined at the teacher-month level, calendar month and calendar year were explicitly incorporated as temporal fixed effects. This approach allowed the model to account for seasonal and academic-calendar patterns, as well as broader disruptions during and after the COVID-19 pandemic. The lower occurrence observed in 2020 may be compatible with reduced interpersonal contact during remote work and school closures, although changes in healthcare access, administrative certification, and reporting practices may also have contributed. The higher odds observed after the return to in-person schooling should therefore be interpreted cautiously, as they may reflect a combination of changes in exposure patterns, healthcare-seeking behaviour, certification practices, and administrative registration over time [30].

Contract type was also associated with sick leave occurrence. Provisional teachers had lower odds of monthly sick leave occurrence than permanent staff (OR 0.619). This finding should not be interpreted as evidence of better health among provisional teachers. It may reflect differences in employment stability, age or career stage, access to certification pathways, perceived job insecurity, or presenteeism, whereby workers avoid taking leave despite illness because of concerns about employment consequences. Further research incorporating measures of job tenure, working conditions, perceived job security, and presenteeism would be needed to clarify the mechanisms underlying this association [31,32].

This study has several strengths. The use of large-scale administrative data allowed comprehensive coverage of the public teaching workforce over an extended period. The teacher-month structure of the main model made it possible to examine monthly occurrence and temporal variation, while annual descriptive summaries allowed the burden of sick leave to be described at the teacher-year level. In addition, the availability of diagnostic information from 2018 onward allowed characterization of the main ICD-10 diagnostic groups associated with general illness–related sick leave.

Several limitations should also be acknowledged. First, reliance on administrative records may introduce limitations related to diagnostic coding accuracy, changes in reporting systems over time, and variation in certification practices. Second, ICD-10 diagnostic information was only available from 2018 onward, which limited long-term diagnostic trend analyses. Third, the databases did not include individual behavioural factors, comorbidities, clinical severity, workplace environmental measurements, classroom density, ventilation, school-level organisational conditions, commuting patterns, or direct measures of psychosocial risk. Fourth, although calendar month and calendar year were included as temporal fixed effects, the observational design does not allow temporal changes to be attributed to specific policy decisions, pandemic-related measures, changes in school operation, or administrative registration practices. Fifth, the primary outcome was defined as a binary indicator of at least one general illness–related sick leave episode in a teacher-month observation. Therefore, the model did not capture the number of episodes within the same month, cumulative days lost, recurrence patterns, or the economic burden of sick leave. Finally, sick leave episodes were linked to calendar months according to the episode start date; therefore, episodes spanning more than one calendar month may not have been fully represented in the monthly binary outcome.

## Conclusion

General illness–related sick leave among public school teachers in Bogotá was a frequent and systematically patterned phenomenon during the study period. Annual descriptive analyses showed a substantial burden of medically certified absence, while the teacher-month multivariable model identified sociodemographic, occupational, territorial, and temporal factors associated with monthly sick leave occurrence. Older age and female sex were associated with higher odds of general illness–related sick leave, and relevant variation was also observed by occupational role, contract type, locality, calendar month, and calendar year.

Respiratory, musculoskeletal, and infectious diseases accounted for the largest share of general illness–related sick leave episodes, whereas chronic, neoplastic, injury-related, circulatory, and mental health conditions were associated with longer absences. These findings provide population-level evidence to support occupational health surveillance, seasonal preparedness, and workforce planning in the public teaching sector. Preventive strategies may benefit from being aligned with academic-calendar and seasonal patterns, while differentiated approaches for musculoskeletal and mental health conditions may help inform more targeted occupational health interventions.

## Data Availability

The data sources for this study comprised three administrative databases provided by the Bogotá District Department of Education (SED) (Additional File 1). Data is available upon request to to SED.

## Acknowledgments

This study is a scientific product derived from the consultancy work carried out between the Universidad Nacional de Colombia and the Secretaría de Educación Distrital de Bogotá under contract: 8188414. The authors express their gratitude to both institutions for the technical support provided during the execution of this project. The funders had no role in study design, data collection and analysis, decision to publish, or preparation of the manuscript.

## Supporting Information

**Additional File 1.** Administrative Database use approval

**Additional File 2.** STROBE Checklist

## Author Contributions

**Conceptualization:** Hernando Bayona-Rodríguez, Daniela Sánchez-Santiesteban, Giancarlo Buitrago.

**Methodology:** Hernando Bayona-Rodríguez, Daniela Sánchez-Santiesteban, Giancarlo Buitrago.

**Data curation:** Daniela Sánchez-Santiesteban, Giancarlo Buitrago.

**Formal analysis:** Daniela Sánchez-Santiesteban, Giancarlo Buitrago.

**Writing – original draft:** Hernando Bayona-Rodríguez, Daniela Sánchez-Santiesteban, Giancarlo Buitrago.

**Writing – review & editing:** Hernando Bayona-Rodríguez, Daniela Sánchez-Santiesteban, Giancarlo Buitrago.

## References

1. Viac C, Fraser P. Teachers’ well-being: A framework for data collection and analysis. 2020 Jan. Report No.: 213. doi:10.1787/c36fc9d3-en

2. Darling-Hammond L. Teacher Quality and Student Achievement. Educ Policy Anal Arch. 2000;8: 1. doi:10.14507/epaa.v8n1.2000

3. Pascual E, Perez-Jover V, Mirambell E, Ivañez G, Terol MC. Job Conditions, Coping and Wellness/Health Outcomes in Spanish Secondary School Teachers. Psychol Health. 2003;18: 511–521. doi:10.1080/0887044031000147238

4. Heinz M, Daid RM, Keane E. The essential role of teacher diversity in creating equitable and inclusive learning environments: an interdisciplinary conceptual framework. Learn Environ Res. 2025;28: 387–407. doi:10.1007/s10984-025-09540-5

5. Levy-Feldman I. The Role of Assessment in Improving Education and Promoting Educational Equity. Educ Sci. 2025;15: 224. doi:10.3390/educsci15020224

6. Akkuş M, Çinkir Ş. The Problem of Student Absenteeism, Its Impact on Educational Environments, and The Evaluation of Current Policies. Int J Psychol Educ Stud. 2022;9: 978–997. doi:10.52380/ijpes.2022.9.4.957

7. Maceke RR, Chauke TA, Nkoana M. Contributing factors to unexcused teacher absenteeism in Adult and Community Education and Training centres. 2025.

8. Madigan DJ, Kim LE. Does teacher burnout affect students? A systematic review of its association with academic achievement and student-reported outcomes. Int J Educ Res. 2021;105: 101714. doi:10.1016/j.ijer.2020.101714

9. Gong X, Yi M, Jiang C, Xiong Q, Xu B, Weng F, et al. Global burden and trends of occupational noise-induced hearing loss (1990-2021) and projection to 2040. Front Public Health. 2025;13: 1682413. doi:10.3389/fpubh.2025.1682413

10. Wang Y, Zhang N, Li X, Du W, Wang H, Shi X. Global health burden and inequality patterns of occupational noise exposure from 1990 to 2019. Sci Rep. 2025;15: 24844. doi:10.1038/s41598-025-09575-x

11. Rogers FH, Vegas E. No More Cutting Class? Reducing Teacher Absence And Providing Incentives For Performance. World Bank; 2009. doi:10.1596/1813-9450-4847

12. UNESCO. Caracterización de los docentes latinoamericanos: Estudio Regional Comparativo y Explicativo (ERCE 2019). 2019.

13. Fuentes-Vilugrón G, Baeza-Vargas Y, Fuentes-Fuentes D, Ortiz-Peña D, Rojas-Estrada D, Sandocal-Obando E, et al. Mental health and burnout levels of early childhood education pedagogical teams in Chile. Front Educ. 2025;10: 1680412. doi:10.3389/feduc.2025.1680412

14. Marenco-Escuderos AD, Ávila-Toscano JH. Burnout y problemas de salud mental en docentes: diferencias según características demográficas y sociolaborales. Psychologia. 2016;10: 91–100. doi:10.21500/19002386.2469

15. Rojas Botero ML, Zapata Hoyos JA, Grisales Romero H. Síndrome de burnout y satisfacción laboral en docentes de una institución de educación superior. Rev Fac Nac Salud Pública. 2009;27: 1–13. doi:10.17533/udea.rfnsp.924

16. Guevara-Manrique AC, Sánchez-Lozano CM, Parra L. Estrés Laboral y Salud Mental en Docentes de Primaria y Secundaria. Revista Colombiana de Salud Ocupaciona. 2014;4: 30–32.

17. Ortiz EG, Rairan DRB, Bustacara EP, Medina DAB, Alvarado BEP. Ausentismo laboral por incapacidad médica en los docentes del magisterio, Distrito de Bogotá. 2015-2019. 2020.

18. Sánchez-Santiesteban D, Bayona-Rodríguez H, Buitrago G. Mortality and comorbidities among teaching professionals: A cross-sectional study in Colombia. Acero A, editor. PLOS One. 2026;21: e0332110. doi:10.1371/journal.pone.0332110

19. STATA. STATA MP. [cited 25 Apr 2023]. Available: https://www.stata.com/statamp/

20. Cuschieri S. The STROBE guidelines. Saudi J Anaesth. 2019;13: S31–S34. doi:10.4103/sja.SJA_543_18

21. Goetzel RZ, Long SR, Ozminkowski RJ, Hawkins K, Wang S, Lynch W. Health, Absence, Disability, and Presenteeism Cost Estimates of Certain Physical and Mental Health Conditions Affecting U.S. Employers: J Occup Environ Med. 2004;46: 398–412. doi:10.1097/01.jom.0000121151.40413.bd

22. Mosley KC, McCarthy CJ, Lambert RG, Fitchett PG, Dillard JB. Elementary teacher occupational health outcomes across schools with varying resources and demographics. Psychol Sch. 2023;60: 943–964. doi:10.1002/pits.22814

23. Guerrero G, Leon J, Zapata M, Cueto S. Getting teachers back to the classroom. A systematic review on what works to improve teacher attendance in developing countries. J Dev Eff. 2013;5: 466–488. doi:10.1080/19439342.2013.864695

24. Ntim SY, Antwi CO, Aboagye MO, Mensah ET, Teye ET, Li X. Emotional labor and absenteeism among early childhood educators: The mediating roles of negative affect and psychological meaningfulness. Heliyon. 2024;10: e40039. doi:10.1016/j.heliyon.2024.e40039

25. De Medeiros AM, Assunção AÁ, Barreto SM. Absenteeism due to voice disorders in female teachers: a public health problem. Int Arch Occup Environ Health. 2012;85: 853–864. doi:10.1007/s00420-011-0729-1

26. Wang, Rui. Are Teachers Absent More? Examining Differences in Absence Between K-12 Teachers and Other College-Educated Workers. 2023 [cited 27 Oct 2025]. doi:10.26300/WERX-6W26

27. Bonfim MMFD, Ferreira LP, Medeiros AMD, Constantini AC, Masson MLV. Voice Disorder, Job Stress, and COVID-19 in Teachers: Impacts in Times of Pandemic. Rev Investig E Innov En Cienc Salud. 2024;6: 8–23. doi:10.46634/riics.231

28. Cezar-Vaz MR, Severo L de O, Borges AM, Bonow CA, Rocha LP. Voice disorders in teachers. Implications for occupational health nursing care. 2012.

29. Ramírez-García CO, Lluguay-Quispillo DJ, Inga-Lafebre JD, Cuenca-Lozano MF, Ojeda-Zambrano RM, Cárdenas-Baque CC. Musculoskeletal Disorders in Primary School Teachers. Sustainability. 2023;15: 16222. doi:10.3390/su152316222

30. Bringing Honesty into Education. September is “Attendance Awareness Month.” In: Notes from the Educational Trenches [Internet]. 11 Sept 2019 [cited 27 Oct 2025]. Available: https://eduhonesty.com/september-is-attendance-awareness-month/

31. Cordoba Calquin CA, Farris M, Rojas Patuelli K. Discussing school socioeconomic segregation in territorial terms: the differentiated influence of urban fragmentation and daily mobility. Investig Geográficas. 2017 [cited 27 Oct 2025]. doi:10.14350/rig.54766

32. Marmot M, World Health Organization, UCL Institute of Health Equity, editors. Review of social determinants and the health divide in the WHO European Region: final report. Copenhagen: World Health Organization, Regional Office for Europe; 2014.

